# COVID-19 in a Designated Infectious Diseases HospitalOutside Hubei Province,China

**DOI:** 10.1101/2020.02.17.20024018

**Authors:** Qingxian Cai, Deliang Huang, Pengcheng Ou, Hong Yu, Zhibin Zhu, Zhang Xia, Yinan Su, Zhenghua Ma, Yiming Zhang, Zhiwei Li, Qing He, Lei Liu, Yang Fu, Jun Chen

## Abstract

**Background:** A new type of novel coronavirus infection (COVID-19) occurred in Wuhan, Hubei Province. Previous investigations reported patients in Wuhan city often progressed into severe or critical and had a high mortality rate.The clinical characteristics of affected patients outside the epicenter of Hubei province are less well understood.

**Methods:** All confirmed COVID-19 case treated in the Third People’s Hospital of Shenzhen,from January 11, 2020 to February 6, 2020, were included in this study. We analyzed the epidemiological and clinical features of these cases to better inform patient management in normal hospital settings.

**Results:** Among the 298 confirmed cases, 233(81.5%) had been to Hubei while 42(14%) had not clear epidemiological history. Only 192(64%) cases presented with fever as initial symptom. The lymphocyte count decreased in 38% patients after admission. The number (percent) of cases classified as non-severe and severe was 240(80.6%) and 58(19.4%) respectively. Thirty-two patients (10.7%) needed ICU care. Compared to the non-severe cases, severe cases were associated with older age, underlying diseases, as well as higher levels of CRP, IL-6 and ESR. The median (IRQ) duration of positive viral test were 14(10-19). Slower clearance of virus was associated with higher risk of progression to severe clinical condition. As of February 14, 2020, 66(22.1%) patients were discharged and the overall mortality rate remains 0.

**Conclusions:** In a designated hospital outside the Hubei Province, COVID-19 patients were mainly characterized by mild symptoms and could be effectively manage by properly using the existing hospital system.

## Introduction

Since December 2019, an outbreak of a new type of novel coronavirusinfection (COVID-19, or 2019-nCoV as previously named) has beenfirst reported in Wuhan (Capital of Hubei Province), China and subsequently spread rapidly to other areas.^1^ Human transmission has been confirmed.^2^ The epidemic has been growing in recent weeks, but the patients linked to the suspected source of transmission, a seafood wholesale market in Wuhan, is decreasing.^3^As of 10:00am, 11/February/2020, there were 42,708 confirmed cases and 1017 deaths in China due to the COVID-19 (National Health Commission). The number of infected persons and death is still increasing very sharply.

Understanding the clinical features of 2019-nCoVinfection is an urgent need to better inform patient management. Recently, Chaolin Huang et al reported a cohort of 41 early emerged patients in Wuhan and concluded the clinical presentations greatly resemble severe acute respiratory syndrome coronavirus (SARS-CoV) and the mortality rate is high (6/41, 15%) in this cohort.^4^ Another report from Wuhan summarized clinical features of 138 hospitalized patients in Wuhan city, presumed hospital-related transmission of 2019-nCoV was suspected in 41% of patients, 26% of patients received ICU care, and mortality was 4.3%.^5^ Besides, Wei-jie Guan et al summarized the Clinical characteristics of 1,099 patients with laboratory-confirmed 2019-nCoV patients from552 hospitals and concluded that severe pneumonia occurred in 15.7%.However, this study included many patients who were treated in local hospitals in Wuhan and their characteristics may not be representative of patients outside of the Wuhan city.^6^It is possible the sudden, clustering onset of 2019-nCoV infection in the Wuhan City has created overwhelming pressures on the local health systems which could influence treatment accessibility and patient prognosis. Additionally, many antiviral drugs targeting RNA virus, including lopinavir/ritonavir, favipiravir, interferon, ribavirin, and abidore/darunavirall, are currently used to treat the COVID-19 on an off-label basis, while their safety and effectiveness in these patients are not fully understood. Here, we investigated 298confirmed COVID-19 cases in a hospital designated to treat all such patients in Shenzhen, Guangdong Province, which is adjacent to the epicenter of the Hubei Province. The number of patients treated in the study hospital has been within the normal range of the hospital capacity.

## Methods

### Study Design and criteria of Participants

Since January 11,allsuspected and confirmed patients with 2019-nCoV were hospitalized in the third people’s Hospital of Shenzhen, which located in Shenzhen, Guangdong Province and is the only hospital authorized to admit the patients diagnosed with 2019-nCoV pneumonia (NCP) by the government in Shenzhen City. The diagnosis of 2019-nCoV infection was based on World Health Organization interim guidance.^1^ The severity of 2019-nCoV acute respiratory disease (ARD) was assessed according to the international guidelines for community-acquired pneumonia.^7^In these studies, we collected all the hospitalized patients data from January 11, 2020 to February 6, 2020 and followed-up to February 11, 2020. The epidemiological history, clinical manifestation, laboratory findings, radiological characteristics, treatment and outcomes data were extracted from electronic medical records.

### Image assess standard

The chest CTs or X-rays were assessed by two specialists and graded to mild, medium and severe change according to the Diagnosis and treatment plan for new coronavirus pneumonia, published by the National Health Commission of the PRC.^8^ A mild CT change is defined by multiple small patchy shadows and interstitial changes mainly involved the outer zone of the lung and under the pleura. Medium changes are defined by ground glass shadows and infiltrative shadows in both lungs. A severe change is defined by diffuse consolidation and uneven density in both lungs. The signs of air bronchus and bronchiectasis can be seen in the non-consolidation area with patchy ground-glass shadow, and “white lung” in most of the affected lungs.

### Real-Time Reverse Transcription Polymerase Chain Reaction Assay for nCoV

The presence of 2019-nCoV was detected by the qPCR method, as previously reported.^5^ Two pairs of primers targeting the open reading frame 1ab (ORF1ab) and the nucleocapsid protein (N) were amplified and examined. The corresponding sequences for ORF1ab were 5’-CCCTGTGGGTTTTACACTTAA-3’ (F), 5’-ACGATTGTGCATCAGCTGA-3’ (R), and 5’-CY3-CCGTCTGCGGTATGTGGAAAGGTTATGG-BHQ1-311 (probe), and those for N were 5’-GGGGAACTTCTCCTGCTAGAAT-3’ (F), 5’-CAGACATTTTGCTCTCAAGCTG-3’ (R), and 5’-FAM-TTGCTGCTGCTTGACAGATT-TAMRA-311 (probe). Each sample was run in triplicate with positive and negative controls set as suggested. These diagnostic criteria were based on the recommendations by the National Centers for Disease Control and Prevention (CDC) of China. Samples identified as positive for 2019-nCoV by the local laboratory were further reconfirmed by the key laboratory of Shenzhen CDC, China. For the confirmed cases, nasal swab samples were collected every 3 days and evaluated by the qPCR assay. The clearance of 2019-nCoV was defined as two consecutive negative results with qPCR detection at an interval of 24 hours.

### Statistical Analysis

Categorical variables were described as frequency rates and percentages, and continuous variables were described using mean, median, and interquartile range (IQR) values. Means for continuous variables were compared using independent group t-tests when the data were normally distributed; otherwise, the Mann-Whitney test was used. Data (no normal distribution) from repeated measures were compared using the generalized linear mixed model. Proportions for categorical variables were compared using the χ^2^ test, although the Fisher exact test was used when the data were limited. Multivariate Cox regression analysis was applied to preliminary evaluate the factors affecting virus clearance. All statistical analyses were performed using SPSS (Statistical Package for the Social Sciences) version 22.0 software (SPSS Inc). For unadjusted comparisons, a 2-sided α of less than .05 was considered statistically significant. The analyses have not been adjusted for multiple comparisons and, given the potential for type I error, the findings should be interpreted as exploratory and descriptive.

### Ethics Approval

Data collection and analysis of cases and close contacts were determined by the National Health Commission of the People’s Republic of China (PRC) to be part of a continuing public health outbreak investigation and were thus considered exempt from institutional review board approval.

### Patient and Public Involvement statement

It was not appropriate or possible to involve patients or the public in the design, or conduct, or reporting, or dissemination plans of our research.

## Results

### Characteristics of 298 patients with 2019-nCoV

As shown in table 1, the median age of all patients was 47 years (IQR, 33-61). 50% of all patients were male. The median BMI was 23.05 (20.92-25.44). 69.1% patients were from Wuhan city, 12.4% from other areas of Hubei province, 4.4% had not been to Hubei provincebut been infected by people from Hubei province;, nad14.1% had no clear contact history; 59.1% was family cluster infection; hospital-related transmission rate occurred in Shenzhen city was 0. A small percentage of patients had pre-existing conditions including diabetes (6.4%), hypertension (12.8%), cardiovascular disease (3.7%), liver diseases (2.7%), malignancy (1.4%), and others (3.7%). The most common symptom at onset of illness was fever (192 [64.4%] patients). 10.1% of the patients have no symptoms at onset.

**Table 1.** Characteristics of 298 patients with 2019-nCoV

|  |  |
| --- | --- |
| Age –yr | 47(33-61) |
| Male/Female - no. | 149/149 |
| BMI | 23.05(20.92-25.44) |
| Epidemic - no. (%) |  |
| From Hubei province | Wuhan 206(69.1),other area of Hubei province 37 ( 12.4 ) |
| Not been to Hubei province, but infected by people from Hubei province | 13(4.4) |
| Without any clear contact history | 42(14.1) |
| Family cluster infection | 176 ( 59.1 ) |
| Hospital-related transmission rate (Shenzhen City) | 0 |
| **Basic disease- no. (%)** |  |
| Diabetes | 19(6.4) |
| Hypertension | 38(12.8) |
| Cardiovascular diseases | 11(3.7) |
| Liver disease | 8(2.7) |
| Malignancy | 4(1.4) |
| Others | 11(3.7) |
| **Initial symptom- no. (%)** |  |
| Fever | 192(64.4) |
| No fever but others | Cough54(18.1),Fatigue6(2.0),Headach4(1.3),Dirrhea6(2.0),sore throat 3(1.0),Nasal congestion 2(0.7) |
| No symptoms | 30(10.1) |
| **Laboratory Examination** |  |
| Blood Routine - Count Median $\times 10^9/L$ (IQR), Abnormal rate(%) | WBC 4.59 ( 3.64-5.63 ) ,3.1;<br>Neutrophils 2.57 ( 1.93-3.49 ) ,3.1;<br>Lymphocyte1.22 ( 0.91-1.74 ) ,38.3 |
| Inflammation factor - Count Median (IQR), Abnormal rate (%) | CRP(mg/dL) 20.91 ( 6.61-47.13 ) ,70;<br>PCT(ug/L) 0.05 ( 0.03-0.08 ) .16.1;<br>IL-6(ng/L) 15.79 ( 7.57-28.72 ) ,76.0;<br>ESR (mm/h) 30 ( 15-50 ) ,60.9 |
| Respiratory index of patients with dyspnea –Median(IQR) | Respiratory rate 27(26-29)<br>Minimum SaO<sub>2</sub> 96(91-95)<br>Minimum oxygenation index 204.5(149.8-265.3) |
| Liver function injury - (Median[IQR],abnormal rate[%]) | Occurrence number and rate 44,14.8<br>ALT (U/L) and abnormal rate 48 ( 24.5-59.5 ) ,8.7<br>AST (U/L) and abnormal rate 48 ( 30-65 ) ,8.7<br>TBIL (umol/L) and abnormal rate 14 ( 10-22 ) ,3.1<br>ALP (U/L)and abnormal rate 63 ( 55.5-70 ) ,3.0<br>GGT (U/L)and abnormal rate44.5 ( 26-66.8 ) ,6.4 |
| Myocardial damage - (Median[IQR],abnormal rate[%]) | Occurrence number and rate 20,6.7<br>CK (U/L) and abnormal rate 283 ( 105-407.5 ) ,3<br>CK-MB (U/L) and abnormal rate 1.5 ( 1.0-4.25 ) ,0<br>LDH (U/L) and abnormal rate 412.5 ( 317.8-687.3 ) ,6.3<br>Myoglobin (ng/mL) and abnormal rate 127.5 ( 67.75-216.25 ) ,3.3 |
| Renal injury - (Median[IQR],abnormal rate[%]) | Occurrence number and rate 17, 5.7<br>BUN (mmol/L) and abnormal rate 8(6-10),4.0<br>Cr (umol/L) and abnormal rate 102(72-122),4,4% |
| D-Dimer - no. rate(%), Median(IQR) | Elevation number and rate 99,35.7<br>0.39 ( 0.27-0.63 ) |
| **Image changes of lung (CT or X-ray) - no. (%)** |  |
| At admission | Mild 119(40.0),Medium170(57.0),Severe9(3.0) |
| **Clinical classification - no. (%)** |  |
| Classification | No-severe 240(80.6), Severe58(19.4) |
| Disease progressive number - no. (%) and days - Median(IQR) | From Onset to Dyspnea 13 ;9(7-11)<br>From Onset to ARDS 13, 9(7-11)<br>From Onset to Severe 58, 9(6.5-11)<br>From Onset to Mechanical ventilation 32, 10(6.8-13.3) |
| Therapy | Antiviral: Lopinavir/ritonavir 229(76.8); Favipiravir 30(10.1)<br>Shorttermmethylprednisolone and human gamma-globulin:all 58 severe patients<br>Antibacterial therapy37(12.4) |
|  | Need ICU32(10.7)<br>Invasive mechanical ventilation 16(5.4)<br>Extracorporeal membrane oxygenation 2(0.6)<br>Kidney replacement therapy4(1.3) |
| Duration from onset to hospital (days) | 5 ( 2-7) |
| Duration from onset to viral clearance of nasal swab (days) | 14 ( 9-19) |
| Hospitalization | In hospital 229, (76.9) Discharge 66,(22.1) |
| Mortality | 0(0) |

Except for percentage, all number shown as Median(IQR); BMI, body mass index; CRP, C-reactive protein;PCT, procalcitonin; IL-6,interleukin-6; ESR,erythrocyte sedimentation rate; ALT,aspartate aminotransferase; AST,utamic oxaloacetic transaminase; TBIL,total bilirubin abnormal; ALP,alkaline phosphatase; GGT,gammaglutamyltranspeptidase; CK,creatine kinase; LDH,lactate dehydrogenase; BUN,urea nitrogen; Cr,creatinine. All number of these indexes showed the peak value of all abnormal patients.

The median of the lymphocyte was 1.22×109/L on admission and decreased in 38.3% patients. The median of inflammation factors of the peak C-reactive protein(CRP) was 20.91 mg/DL (6.61-47.13)and increased in 70% patients, peak procalcitonin (PCT) was 0.05 ug/L(0.03-0.08) and increased in 70% patients, peak interleukin-6 (IL-6) was 15.79 ng/L(7.57-28.72) and increased in 76% patients, and the peak erythrocyte sedimentation rate (ESR) was 30 mm/h (15-50) and increased in 60.9% patients. The respiratory index of patients with dyspnea of respiratory rate was 27(26-29), and the minimum oxygenation index was 204.5(149.8-265.3). The occurrence number and rate of liver injury were 44(14.8%). The myocardial damage number and rate was 20(6.7%). The occurrence rate of renal injury was 5.7%. D-Dimer was increased in 99 cases (35.7%). Chest CT or X-ray images of the 298 patients, 119 (40%) was mild, 170(57%) was medium and 9(3%) was severe on admission. Figure 1 is a typical patient’s imaging change from mild to severe.

**Fig 1.**
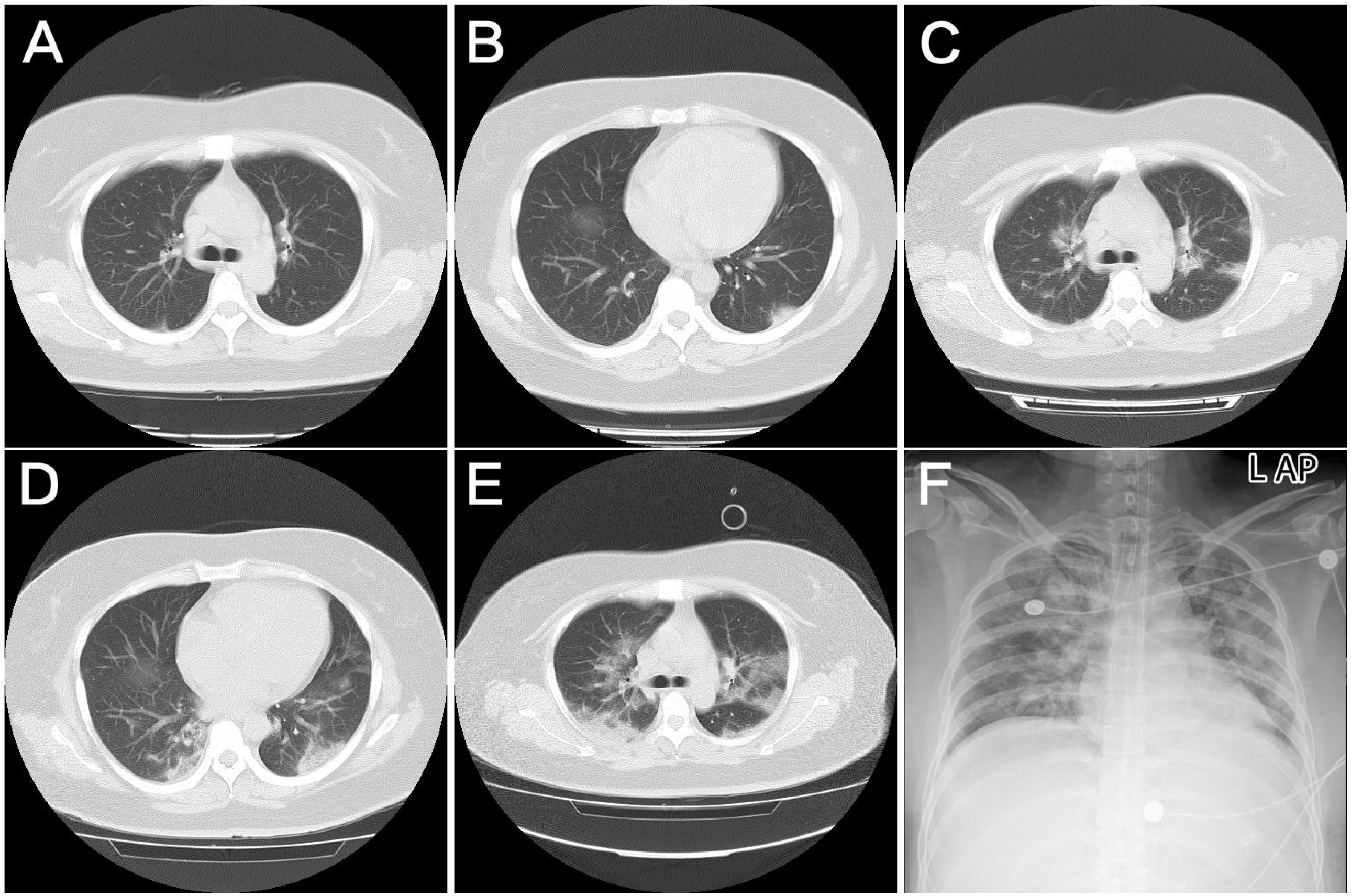
The Imageology change with the disease progression of a 36 years-old female severe patient infected with 2019-CoV. A and B were second day of the course of disease; C and D were day 7; E was day 11 and F was day 15. A. CT showed local ground glass shadow and nodules; B. Thin ground glass shadow Mainly distributed in outer zone of the lung and under the pleura; C. Ground glass shadow of both lungs, left pulmonary nodule focus located under pleura; D. Progress to patchy shadow with uneven density; E. Multiple patchy shadows with fuzzy edges were observed, and large ground glass shadow and condensation shadow were found in both lower lobe; F. Chest radiograph showed the brightness of both lungs was decreased, “white lung” in all over most of the lungs.

**Fig 2.**
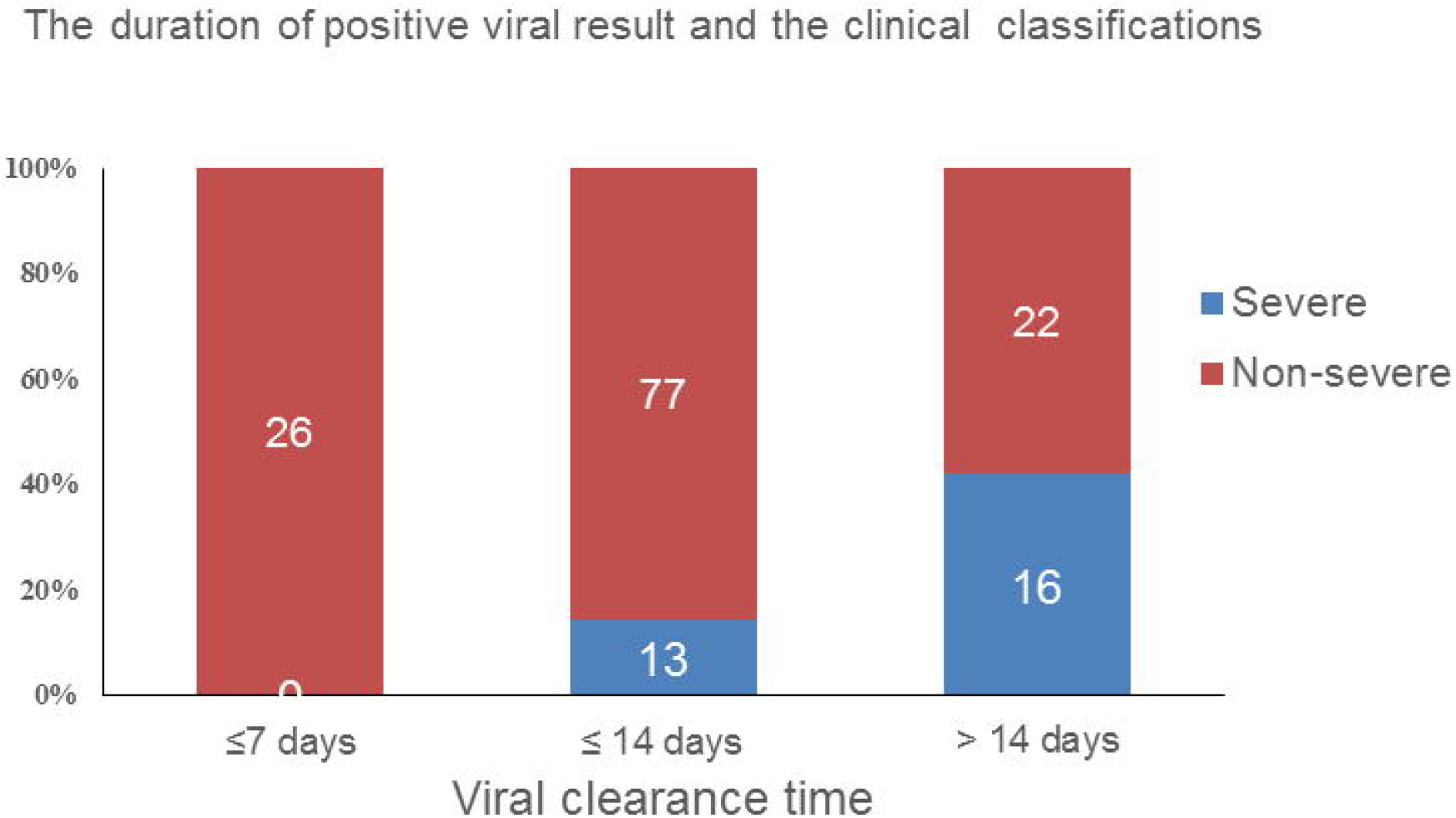
A total of 175 cases reach the endpoint of viral clearance. The chi square test shown the distribution of severe clinical condition was different among the patients with different duration of positive viral test result (P<0.001). Of the patients with virus clearance within 7 days from disease onset, no one progress to severe cases. Of those with viral clearance within 14 days from disease onset, 13/90(14.4%) patients progress to severe cases. Of those still had positive viral test after 14 days from disease onset, 30/71(42.3%) patients progress to severe cases.

In our patients, 240(80.6) and 58(19.4) patients were categorized into non-severe and severe subgroups, respectively. During hospitalization, the number of days betweenillness onset anda serious complication was 9 (7-11) for dyspnea, 9(7-11) for ARDS, 9(6.5-11) for severe, and 10(6.8- 13.3) for mechanical ventilation. The average duration from onset to hospitalization was 5(2-7) days. As of February 11, 2019, 66(22.1%) of the 298 patients hadmet the discharged criteria and no patients died.

Most patients received antiviral therapy, with 76.8% receiving Lopinavir/ritonavir, and 10.1% receiving Favipiravir. For all severe cases, 3-5 days duration of intravenous methylprednisolone (1- 2mg/kg/d) combined with human gamma-globulin (10-20g per day) were prescribed. 37 patients received antibacterial therapy. 16 cases received invasive mechanical ventilation, of which 2 patients received extracorporeal membrane oxygenation as rescue therapy. 4 patients received renal dialysistherapy.

### Comparison of severe and non-severe patients with 2019-nCoV

Patient age differed significantly between the two subgroups (Table 2; 40.0 *vs*. 56.0 years, p<0.001). Compared with non-severe cases, the underlying medical conditions were significantly more common in severe groups. The duration of the fever is marked longer in severe group (p=0.024).

**Table 2.** Comparison of different classification patients with 2019-nCoV.

|  | **Non-Severe**<br>**N=240** | **Severe**<br>**N=58** | ***p value*** |
| --- | --- | --- | --- |
| Age (yr) | 40.0 ( 31.0-56.0 ) | 64.0 ( 56.0-66.0 ) | <0.001 |
| Male sex - no. (%) | 111/240(46.3) | 33/58(56.9) | 0.145 |
| BMI | 22.86 ( 20.5-25.2 ) | 23.86 ( 22.0-26.7 ) | 0.353 |
| Basic diseases - no. (%) | 60(25.0) | 36(62.1) | <0.001 |
| Duration of fever - no. (%) | 5 ( 3-8 ) | 6 ( 4-8 ) | 0.024 |
| peak temperature - no. (%) | 38 ( 37.8-38.8 ) | 38.95 ( 38.5-39.2 ) | 0.063 |
| Lymphocyte count($\times 10^9/L$) – median(IQR) | 1.3 ( 1.0-1.8 ) | 0.97 ( 0.65-1.19 ) | <0.001 |
| Lymphocyte<1.1*1 - no. (%) | 75(31.3) | 39(67.2) | <0.001 |
| CRP(mg/dL) – median (IQR) | 14.24 ( 4-34.38 ) | 62.9 ( 32.99-110.67 ) | <0.001 |
| CRP>8 U/L - no. (%) | 142(59.2) | 54(93.1) | <0.001 |
| IL-6(ng/L) – median(IQR) | 12.0(6.4-19.7) | 38.8(22.7-57.2) | <0.001 |
| IL-6>7 - no. (%) | 110(45.8) | 36(62.1) | 0.026 |
| ESR (mm/h) – median (IQR) | 25(14-42.3) | 49(39.5-62.5) | <0.001 |
| ESR> 20mm/h - no. (%) | 82(34.2) | 35(60.3) | <0.001 |
| D-Dimer ( mg/L ) – median (IQR) | 0.35(0.25-0.52) | 0.96(0.54-1.925) | <0.001 |
| D-Dimer>0.5mg/L - no. (%) | 60(25.0) | 39(67.2) | <0.001 |
| CK>200U/L - no. (%) | 3(1.3) | 6(10.3) | <0.001 |
| LDH>250 U/L - no. (%) | 4(1.7) | 15(25.9) | <0.001 |
| Myoglobin>110 U/L - no. (%) | 1(0.4) | 9(15.5) | <0.001 |
| Percentage of liver injury patients - no. (%) | 23 ( 9.6 ) | 21 ( 36.2 ) | <0.001 |
| Percentage of renal injury patients - no. (%) | 4 ( 1.7 ) | 13 ( 22.4 ) | <0.001 |
| Percentage of myocardial damage | 5(2.1) | 15(25.9) | <0.001 |
| - no. (%) |  |  |  |
| Duration from onset to hospital (days) - no. (%) | 4.5(2.0-7.0) | 5(4.0-6.8) | 0.071 |

Severe patients had lower lymphocyte counts than non-severe patients. The median of inflammation factors of the CRP, IL-6 and ESR was much higher in the severe group, similarly in the proportion of CRP>8, IL-6>7, and ESR> 20mm/h patients. The level of D-dimer on admission was elevated markedly in the severe group compared with non-severe (median 0.35mg/L [0.25- 0.52], p<0.001), and the number of D-dimer>0.5mg/L in the severe group are significantly more than the other. Concerning the myocardial damage, the quantity of myocardial damage of CK>200U/L, LDH>250U/L and myoglobin>110 U/L in the severe group is larger than the other patients. The percentage of liver injury, renal injury and myocardial damage patients was also greatly enhanced in the severe group compared with the non-severe group.

### Factors affecting virus clearance

The duration of positive viral test resultwas defined as the time from the day of disease onset to the day of virus clearance, which was defined as two consecutive negative results with qPCR detection at an interval of 24 hours(using 1st day).As shown in table 3, patient age(47 years orolder*vs*. below), clinical classification(severe *vs*.non-severe)were independent prognostic factors of viral clearance with adjustedhazard ratios of 0.56 (95% confidence interval 0.40-0.76, *p*< 0.001). Chi-square test found the rates of patients progress to severe clinical condition was correlated with duration of positive viral test result(P<0.001), as shown in figure2.

**Table 3.** The factors affecting duration of positive viral test result

|  |  |  | Kaplan-Meier analysis |  | Cox regression models |  |
| --- | --- | --- | --- | --- | --- | --- |
| Risk factor |  | N | Duration(days)<br>Median[IQR] | P | Adjusted hazard ratio | P |
| Total | — | 298 | 14(10-19) |  |  |  |
| Age | ≤47 | 150 | 12(8-16) |  |  |  |
|  | >47 | 148 | 16(11-21) | <0.001 | 0.84(0.57-1.24) | 0.37 |
| Gender | Female | 149 | 12(10-17) |  |  |  |
|  | Male | 149 | 15(10-21) | 0.05 | 1.13(0.78-1.64) | 0.52 |
| BMI | 23.9 | 183 | 13(9-18) |  |  |  |
|  | ≥24 | 115 | 13(10-21) | 0.12 | 0.67(0.45-0.98) | 0.04 |
| Clinical Classification | Non-severe | 240 | 18(14-24) |  |  |  |
|  | Severe | 58 | 13(9-16) | 0.05 | 0.62(0.38-1.00) | 0.05 |
| Antiviral therapy | No | 39 | 14(10-19) |  |  |  |
|  | Yes | 259 | 15(10-19) | 0.446 | 0.62(0.31-1.22) | 0.17 |

BMI, body mass index.

## Discussion

Besides SARS-CoV (outbreak in 2002) and Middle East respiratory syndrome coronavirus (MERS-CoV) (outbreak in 2012), 2019-nCoV is the third coronavirus to emerge in the human population that has put global public health institutions on high alert. Current studies confirmed 2019-nCoV is a novel beta-coronavirus belonging to the sarbecovirus subgenus of Coronaviridae family and has more than 85% identity with a bat SARS-like CoV genome published previously.^9^

The spread of the 2019-nCoV infections in China was much faster than that of the 2002 SARS (over 30,000 cases in 2 months*vs*. 4,698 cases in 6 months). Some epidemic prediction models also showed the basic reproductive number (R_0_) of the 2019-nCoVwas much higher than SARS.^2, 3^ In our study, most of the cases were from Hubei province (81.5%), especially from Wuhan city (69.1%). 14.8% had not been to Hubei province, but infected by people from Hubei province. Family cluster infection is very common (59.1%). Thisclearly demonstrates the importance of disease control measures to prevent imported cases and transmission within close contacts. Moreover, there were 42 cases without any clear contact history, which may indicate third generation cases become emerging in Shenzhen city. Controlling the growth of those cases could be a very tough challenge for other areas out of Hubei Province.

Our data demonstrated older people tended to develop to severe or critical conditions. Severe or critical patients with basic diseases were more common than without basic diseases (62.1% *vs*. 25.0%, *p*<0.001). Therefore, access to high quality medical treatments should be prioritized for this patient group. BMI seems to have little effect on the progress of the disease.

For SARS patients, fever is almost always presented as first clinical manifestations.^10^ However, our data showed more than 1/3(106) 2019-nCoVpatients had no fever at initial time of disease onset. Further, in those 106 patients without fever in initial time, 47 patients never had fever throughout the entire disease course. This suggested limitations in using body temperature alone to screen the disease. Additional indications should be included in the screening criteria.The duration of fever in severe patients was much longer than mild patients (median 6[4-8] *vs*. 5[3-8], *p*<0.024), peak temperature in the severe or critical patients tends higher than mild patients but had no significant difference (*p*=0.063).

Inflammatory responses triggered by viral infection play a crucialrole in pulmonary pathology severity.^11^ Our data confirmed the existence of an inflammatory factor storm; inflammatory factors increased in most of the patients, especially for CRP increased in 70% patients, IL-6 in 76.0% patients, ESR in 60.9% patients. Furthermore, compared to no-severe patients, the above three factors significant increased among severe patients (*p*<0.001). That suggested the three inflammatory factors may help judge the disease progression. Moreover, suppressing the hyperintense immune response to reduce lung inflammation may be a valuable treatment method, such as glucocorticoid and gamma globulin. Glucocorticoids often cause severe secondary infections; the application timing, duration and dosage need further investigation.

Except for respiratory track damage, other organ damagesare of concern for the COVID-19 patients. Our data showed liver injury seldom occurred in these patients (44/298, 14.8%), and mainly in severe patients (36.2% *vs*. 9.6%, *p*<0.001). 55.4% of liver injuries occurred following administration of lopinavir and ritonavir,suggesting possible drug-induced liver injury. Previous *in vitro* experiments revealed that 2019-nCoV might directly bind to ACE2 positive cholangiocytes but not necessarily hepatocytes.^12^ Our data showed the characteristic of the liver injury was hepatocytes related enzymes ALT and AST elevated slightly. Cholangiocytes related enzymes AKP and GGT also elevated in a few patients (3.1% and 3.0%) and elevated slightly. Myocardial damage is relatively rare (6.7%), mainly happened in severe patients (25.9% *vs*. 2.1%, *p*<0.001). Most of the renal injury also happened in severe patients (22.4% *vs*. 1.7, *p*<0.001), the median of BUN and Cr just slightly elevated (8[6-10] mmol/L and 102[72-122] umol/L). All these organ damages may be attributed to the complicated pathophysiological process at the advanced disease stage. We could not conclude that the organ injurywas directly related to the virus infection. Moreover, our data showed 8-10 days of hospitalization is a crucial time point for patients progressing into severe respiratory outcomes, including dyspnea, ARDS, from no-severe to severe and need mechanical ventilation.

In previous reportsof COVID-19 in the Wuhan city,^3, 4^nearly 1/3 patients were admitted to the ICU. The hospital-related transmission rate and mortality was high. This may reflect the very tense situation in Wuhan city, the number of patients is too large and crowded in the hospital, the waiting time for hospitalization is too long, far exceeding the hospital load which caused the above results.However, the severity as shown in Figure 1 is very rare in our hospital. Our hospital data showed there just 11.4% were critical patients who need ICU care, most of the patients presented no-severe (80.6%), hospital-related transmission rate and mortality keep zero. That indicated 2019-nCoV may be less pathogenic, the rate of progressed to severe or critical cases is not so frequency as SARS or MERS (range from 70 to 90%),^13-15^ properly running of the hospital system and can provide timely and effective treatment for patients, greatly reduce hospital-related transmission rate and mortality rate.

Antiviral drugs specifically targeting 2019-nCoV is in developing but the process could be slow. Remdesivir was reported might effective in one patient,^16^ but only one case. In our hospital, lopinavir/ritonavir and favipiravir were widely used, our multivariate Cox regression analysis results showed age and severe type were identified as an independent prognostic factor. However, gender,BMI and antiviral agents lopinavir/ritonavir or favipiravir were not independent prognostic factor for virus clearance. As there were too many confounding factors, the efficacy of the drug needs to be further confirmed by randomized controlled trials(RCT). The use of antibiotics in these patients is controversial, in our hospital, antibacterial therapy was strictly monitored and prescribed only to those with a confirmed bacterial infection or some highly suspected cases, except for those underwent mechanical ventilation or otherwise indicated.

In conclusion, our data demonstrated imported cases are the majority of COVID-19 in the study hospital.The elder and those having basic diseases were more likely progress to severe conditions. Fever at an initial time only presented at 64.4% cases.Liver, renal and myocardial injury seldom happened and most of them happened at the end age patients. Lymphocyte decrease in blood routine,inflammation factor CRP,IL-6 and ESR increase, D-Dimer increase may take as characteristics of laboratory examination. Most of the cases presented as mild, and severe cases were less frequent compared to SARS or MERS. For patients progressing into severe respiratory outcomes, deterioration usually occurred 6-10 days after disease onset. Properly running of hospital system may greatly reduce hospital-related transmission rate and mortality rate. The effect of off-label use antiviral drugs needs more evidence by RCT or real-world studies.

## Data Availability

The data that support the findings of this study are available from the corresponding author on reasonable request. After the publication of the study findings, the data will be available for others to request. The research team will provide an email address for communication once the data are approved to be shared with others. The proposal with a detailed description of study objectives and statistical analysis plan will be needed for evaluation of the reasonability to request for our data. The corresponding author will make a decision based on these materials. Additional materials may also be required during the process.

## Acknowledgments

This work is funded by the National infectious diseases Clinical Research Center.

## Contributors

JC and LL had the idea for and designed the study and had full access to all data in the study and take responsibility for the integrity of the data and the accuracy of the data analysis. QC, DH, PO, MZ, YZ and YF contributed to the writing of the report. HY, ZZ and QH contributed to the critical revision of the report. QC, ZX,YS and ZL contributed to thestatistical analysis. All authors contributed to data acquisition, data analysis, or data interpretation, and reviewed and approved the final version.

